# ddPCR: a more sensitive and accurate tool for SARS-CoV-2 detection in low viral load specimens

**DOI:** 10.1101/2020.02.29.20029439

**Authors:** Tao Suo, Xinjin Liu, Jiangpeng Feng, Ming Guo, Wenjia Hu, Dong Guo, Hafiz Ullah, Yang Yang, Qiuhan Zhang, Xin Wang, Muhanmmad Sajid, Zhixiang Huang, Liping Deng, Tielong Chen, Fang Liu, Xu Ke, Yuan Liu, Qi Zhang, Yingle Liu, Yong Xiong, Guozhong Chen, Ke Lan, Yu Chen

## Abstract

Real time fluorescent quantitative PCR (RT-PCR) is widely used as the gold standard for clinical detection of SARS-CoV-2. However, due to the low viral load in patient throats and the limitations of RT-PCR, significant numbers of false negative reports are inevitable, which results in failure to timely diagnose, early treat, cut off transmission, and assess discharge criteria. To improve this situation, an optimized droplet digital PCR (ddPCR) was used for detection of SARS-CoV-2, which showed that the limit of detection of ddPCR is significantly lower than that of RT-PCR. We further explored the feasibility of ddPCR to detect SARS-CoV-2 nucleic acid from 77 clinical throat swab samples, including 63 suspected outpatients with fever and 14 supposed convalescents who were about to discharge after treatment, and compared with RT-PCR in terms of the diagnostic accuracy. In this double-blind study, we tested, surveyed subsequently and statistically analyzed 77 clinical samples. According to our study, 26 samples from COVID-19 patients with RT-PCR negative were detected as positive by ddPCR. No FPRs of RT-PCR and ddPCR were observed. The sensitivity, specificity, PPV, NPV, NLR and accuracy were improved from 40% (95% CI: 27–55%), 100% (95% CI: 54–100%), 100%, 16% (95% CI: 13–19%), 0.6 (95% CI: 0.48–0.75) and 47% (95% CI: 33–60%) for RT-PCR to 94% (95% CI: 83–99%), 100% (95% CI: 48–100%), 100%, 63% (95% CI: 36–83%), 0.06 (95% CI: 0.02–0.18) and 95% (95% CI: 84–99%) for ddPCR, respectively. Moreover, 14 (42.9 %) convalescents still carry detectable SARS-CoV-2 after discharge. Overall, ddPCR shows superiority for clinical diagnosis of SARS-CoV-2 to reduce the false negative reports, which could be a powerful complement to the current standard RT-PCR. It also suggests that the current clinical practice that the convalescent after discharge continues to be quarantined for at least 2 weeks is completely necessary which can prevent potential viral transmission.

## Introduction

The pandemic of coronavirus disease 2019 (COVID-19) caused by the infection of severe acute respiratory syndrome coronavirus 2 (SARS-CoV-2, also refers as HCOV-19) poses a great threat to public health worldwide [1,2]. It presents a huge challenge for the diagnosis of this pathogen. According to World Health Organization (WHO) and Chinese Center for Disease Control and Prevention (CDC), the current gold standard for the diagnosis of SARS-CoV-2 infection is based on the real-time fluorescent quantitative PCR (RT-PCR), which can detect the nucleic acid of SARS-CoV-2 from patient’s specimen [3,4]. The advantages of RT-PCR method are high throughput and relatively sensitive. However, it has been found in clinical practice that some patients had fever and showed symptoms of suspected viral pneumonia such as lower lobe lesions of the lungs by chest computed tomography (CT), but the nucleic acid test of throat swab using RT-PCR did not show positive results until 5–6 days after the onset of viral pneumonia. Remarkably, it was reported that around 60% of SARS-CoV-2 infections are asymptomatic [5]. Moreover, it was estimated that only 30%-60% positive results can be obtained among COVID-19 patients that further confirmed by chest CT and other diagnostic aid [6]. This might be explained by the relatively low viral load in the throat swabs of patients and the sensitivity limitation of RT-PCR technology, which inevitably produced the false negatives during the clinical diagnosis, leading to a potential risk of viral transmission. Moreover, supposed convalescents, who are about to discharge, especially need viral nucleic acid test with true negative results for confirmation and the out of quarantine to avoid virus transmission and recurrence. Therefore, it is an urgent need for a more sensitive and accurate detection method for the pathogenic detection complementary to current ones.

Digital PCR is based on the principles of limited dilution, end-point PCR, and Poisson statistics, with absolute quantification as its heart [7]. It has broader dynamic range without external interference and robustness to variations in PCR efficiency [8–10]. In 2011, Hindson developed the droplet digital PCR (ddPCR) technology based on traditional digital PCR [11]. The reaction mixture can be divided into tens of thousands of nanodroplets during the process. These vast and highly consistent oil droplets substantially improve the detection dynamic range and accuracy of digital PCR in a low-cost and practical format [12]. In recent years, this technology has been widely used, such as analysis of absolute viral load from clinical samples, analysis of gene copy number variation, rare allele detection, gene expression, microRNA analysis and genome edit detection *et al* [13–16].

To improve the diagnostic accuracy of nucleic acid detection of SARS-Cov-2 in low viral load samples using droplet digital PCR, we compared the dynamic range and the limit of detection (LoD) with a 95% repeatable probability between ddPCR and RT-PCR in laboratory, and tested the clinical samples from 77 patients by both ddPCR and RT-PCR for head to head comparison.

## Materials and methods

### Ethics statement

The institutional review board of Renmin Hospital of Wuhan University approved this study (WDRY2020-K089). Written informed consents were obtained.

### Specimen collection and RNA extraction

Data collection was planned before the index test and reference standard were performed (prospective study). To perform the tests of clinical samples, throat swab samples of 63 suspected outpatients and 14 supposed convalescents were randomly collected by the medical staffs from COVID-19 designated Renmin and Zhongnan Hospital of Wuhan University. Throat swab samples of each patients were firstly collected for official approved RT-PCR diagnosis in hospitals and blinding laboratory RT-PCR and ddPCR tests simultaneously with the same primers/probe sets approved by Chinese CDC. The patients were conducted by hospitals independently. All the events happened in hospitals and laboratories are blinded to each other during the tests. The follow-up survey and clinical information of enrolled cohort were collected after the laboratory tests by the medical staffs.

### Specimen collection and RNA extraction

To perform the tests of clinical samples, throat swab samples of 63 suspected outpatients and 14 supposed convalescents were randomly collected by the medical staffs from COVID-19 designated Renmin and Zhongnan Hospital of Wuhan University. Throat swab samples were collected via mouth according to the interim guidance of WHO, and soaked in 500 μl PBS and vortexed with diameter of 3 mm beads (Novastar, China) for 15 seconds immediately. Total RNA was extracted from the supernatant using QIAamp viral RNA mini kit (Qiagen) following manufacturer’s instruction. First strand cDNA was synthesized using PrimeScript RT Master Mix (TakaRa) with random primer and oligo dT primer for subsequent tests of both RT-PCR and ddPCR in laboratory simultaneously.

### Primers and probes

The primers and probes (RainSure Scientific) target the ORF1ab and N of SARS-CoV-2 according to Chinese CDC.

Target 1 (ORF1ab), forward: 5'-CCCTGTGGGTTTTACACTTAA-3',

reverse: 5'-ACGATTGTGCATCAGCTGA-3',

probe: 5'-FAM-CCGTCTGCGGTATGTGGAAAGGTTATGG-BHQ1–3';

Target 2 (N), forward: 5'-GGGGAACTTCTCCTGCTAGAAT-3',

reverse: 5'-CAGACATTTTGCTCTCAAGCTG-3',

probe: 5'-HEX-TTGCTGCTGCTTGACAGATT-TAMRA-3' [17].

### Droplet Digital PCR workflow

All the procedures follow the manufacture instructions of the QX200 Droplet Digital PCR System using supermix for probe (no dUTP) (Bio-Rad). Briefly, the TaqMan PCR reaction mixture was assembled from 2× supermix for probe (no dUTP) (Bio-Rad), 20× primers and probe mix (final concentrations of 900 nM and 250 nM, respectively) and template (variable volume, cDNA of clinic sample or linear DNA standard) in a final volume of 20 μl. Twenty microliters of each reaction mix was converted to droplets with the QX200 droplet generator (Bio-Rad). Droplet-partitioned samples were then transferred to a 96-well plate, sealed and cycled in a T100 Thermal Cycler (Bio-Rad) under the following cycling protocol: 95°C for 10 min (DNA polymerase activation), followed by 40 cycles of 94°C for 30 s (denaturation) and 60°C for 1 min (annealing) followed by an infinite 4-degree hold. The cycled plate was then transferred and read in the FAM and HEX channels using the QX200 reader (Bio-Rad). To avoid the risk of viral infection and false positive results potentially due to the laboratory contamination, all the experiments were done inside the biosafety cabinet in negative pressure biosafety laboratory using filter tips.

### RT-PCR

The primers and probes used in ddPCR are also used in the RT-PCR system that established in laboratory. A 20 μl reaction mix was set up containing 8 μl of template (variable volume, cDNA of clinic sample or linear DNA standard), 10 μl of reaction buffer, 1 μl 20× primers and probe mix and 1 µl Platinum Taq DNA Polymerase mix (Thermo fisher). Thermal cycling was performed at 95°C for 5 min and then 40 cycles of 95°C for 10s, 55°C for 40 s in BIO-RAD CFX96 Touch Real-Time PCR Detection system (Bio-Rad). To avoid the risk of viral infection and laboratory contamination, the same biosafety measures were taken as that for ddPCR.

### Dynamic range and LoD of RT-PCR and ddPCR

The linear dynamic range of the RT-PCR and ddPCR assay was assessed using the serial dilutions of the linear DNA standard containing the target region. To determine the LoD of both RT-PCR and ddPCR, cDNA of throat swab samples of healthy people was spiked with the linear DNA standard in serial concentrations close to the detection limits. The LoD was calculated by Probit regression analysis with a 95% repeatable probability, which is a commonly used type of regression analysis when empirically determining the limit of analyte that can be reliably detected by molecular assays [18].

### Data statistical analysis

Analysis of the ddPCR data was performed with Quanta Soft analysis software v.1.7.4.0917 (Bio-Rad) to calculate the concentration of the target. The positive populations for each primer/probe are identified using positive and negative controls with single (i.e., not multiplexed) primer–probe sets. In addition, plots of linear regression were conducted with GraphPad Prism 7.00, and probit analysis for LoD was conducted with MedCalc software v19.2.1.

The cases of lost contact were not included in our analysis study due to the unclear conditions. The suspected reports of ddPCR (need further detection) were not included in our analysis study according to the ethics statement as no more sampling for laboratory tests. The detection results were compared to the follow-up survey including clinical records of chest computed tomography, IgM/IgG, other etiological detection, and further official approved RT-PCR confirmation 2-12 days later *et al*. The diagnostic performance of RT-PCR and ddPCR were calculated by MEDCALC (https://www.medcalc.org/calc/diagnostic_test.php).

## Results

### Comparison of the dynamic range of ddPCR and RT-PCR

To compare the dynamic range of ddPCR and RT-PCR, serial dilutions of a positive control linear DNA standard of SARS-CoV-2 were tested using primers/probe sets targeting ORF1ab and N of SARS-CoV-2 for both ddPCR and RT-PCR. As shown in Figure 1, the reportable range of ddPCR is 10 to 5×10^4^ copies/reaction for both ORF1ab and N primes/probe sets with R2 = 0.9935 and 0.9908, respectively (Figure 1(A, B)). Meanwhile, those of RT-PCR is 1000 to 10^7^ copies/reaction for both ORF1ab and N primes/probe sets with R2 = 0.9921 and 0.9898, respectively (Figure 1(C, D)). The results showed that the minimum detection range of ddPCR is significantly lower than that of RT-PCR.

**Fig. 1.**
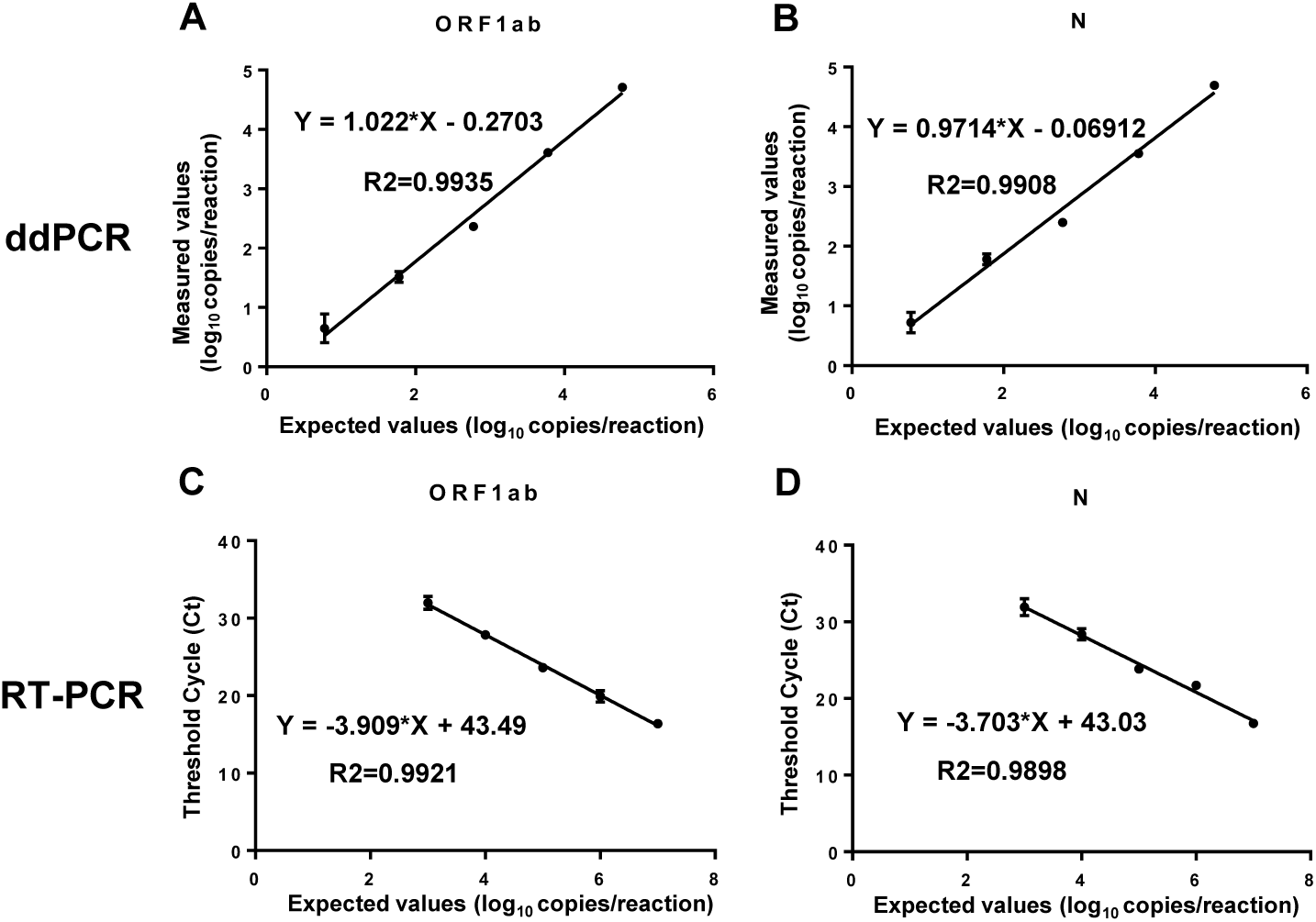
Plot of results from a linearity experiment to determine the reportable range of ddPCR and RT-PCR targeting ORF1ab and N of SARS-CoV-2. (A and B) Expected values (converted to log_10_) were plotted on the X axis versus measured values of ddPCR (converted to log_10_) on the Y axis using Graph Pad Prism targeting (A) ORF1ab and (B) N. (C and D) Expected values (converted to log_10_) were plotted on the X axis versus measured Ct values of RT-PCR on the Y axis using Graph Pad Prism targeting (C) ORF1ab and (D) N. Data are representative of three independent experiments with 3 replicates for each concentration (means ± SD).

### Comparison of the th LoD between ddPCR and RT-PCR

To further determine the accurate LoD of ddPCR and RT-PCR, a series linear DNA standard were diluted to the concentrations below the minimum detection range of ddPCR or RT-PCR by the cDNA of throat swab samples from healthy people (with negative serum SARS-CoV-2 IgM/IgG), which could benefit to reduce the false positive result (FPR). Each concentration was analyzed with 8 replicates. The LoD was calculated by probit regression with a 95% repeatable probability. As shown in Figure 2, the LoD (95 % probability) of ddPCR are 2.1 (95% CI: 1.5-4.2) copies/reaction and 1.8 (95% CI: 1.4-3.3) copies/reaction for ORF1ab (Figure 2(A)) and N (Figure 2(B)) primers/probe sets, respectively. In contrast, the LoD (95% probability) of RT-PCR are 1039 (95% CI: 763.2-1862) copies/reaction and 873.2 (95% CI: 639.8-1633.2) copies/reaction for ORF1ab (Figure 2(C)) and N (Figure 2(D)) primers/probe sets, respectively. Taken together, with the same ORF1ab and N primes/probe sets and template, ddPCR for SARS-CoV-2 detection with a 95% probability, is around 500 times more sensitive than RT-PCR in low level analyte.

**Fig. 2.**
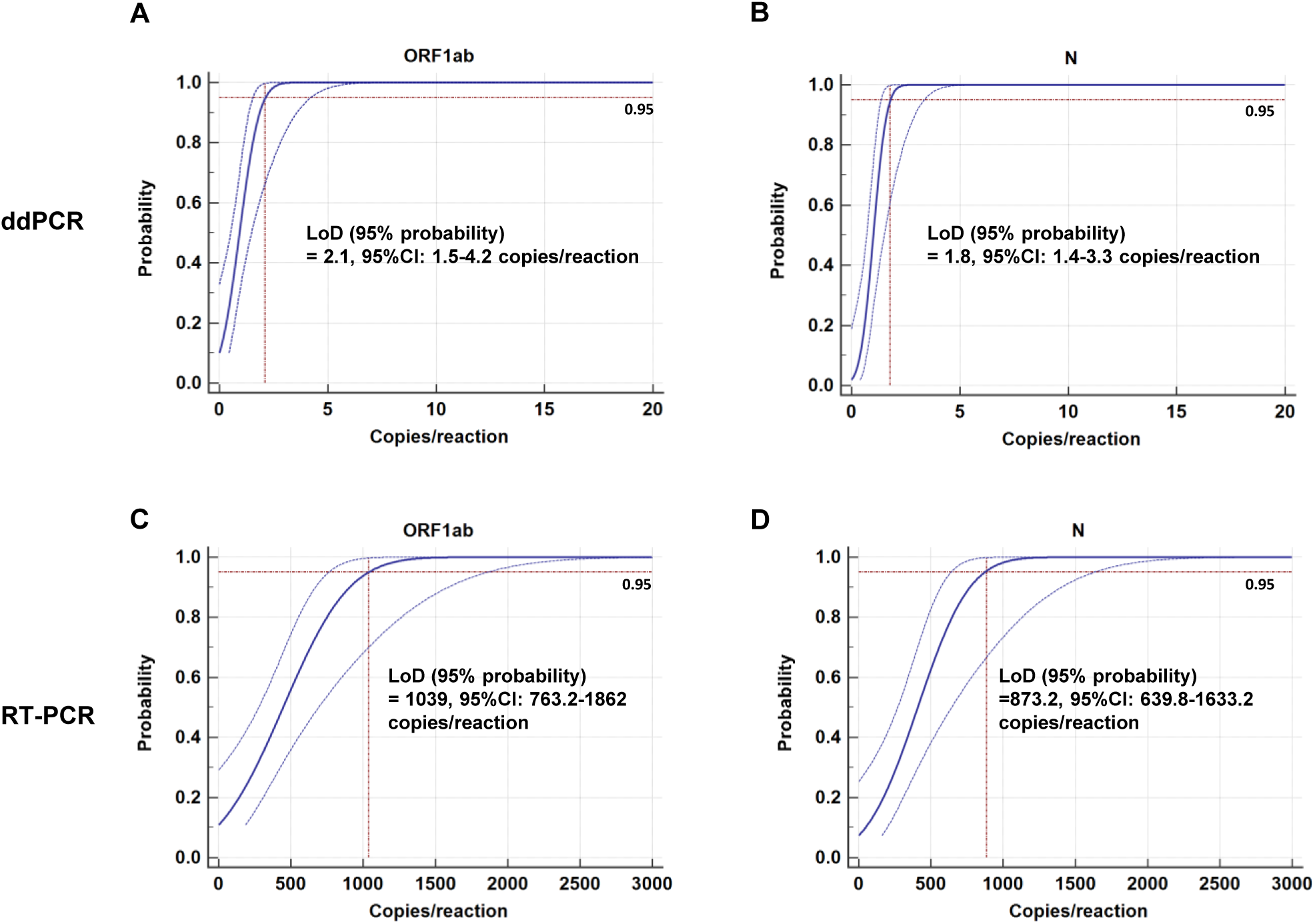
Probit analysis sigmoid curve reporting the LoD of ddPCR and RT-PCR. Replicate reactions of (A) ORF1ab and (B) N of ddPCR or (C) ORF1ab and (D) N of RT-PCR were done at concentrations around the detection end point determined in preliminary dilution experiments. The X axis shows expected concentration (copies/reaction). The Y axis shows fraction of positive results in all parallel reactions performed. The inner line is a probit curve (dose-response rule). The outer lines are 95% confidence interval (95% CI). Data are representative of three independent experiments with 8 replicates for each concentration.

### Detection of SARS-CoV-2 nucleic acid from patient throat swab samples with ddPCR and RT-PCR

To avoid the FPR potentially due to the laboratory contamination, all the experiments were performed inside the biosafety cabinet in negative pressure biosafety laboratory using filter tips. To further assess the systematic FPR, the cDNA of throat swab samples from healthy people were tested with ddPCR for 32 repeats (16 repeats for each primers/probe set, data not shown). In total, 29 out of 32 results (91 %) showed negative read out (0 copy/reaction). 3 out of 32 results (9 %) showed one single positive droplet (around 1 copy/reaction), which is less than the LoD of ddPCR for ORF1ab (2.1 copies/reaction) and N (1.8 copies/reaction) primers/probe sets. Therefore, the positive threshold of ddPCR for SARS-CoV-2 detection is defined as equal as or greater than the LoD of ddPCR for ORF1ab and N primers/probe sets, respectively. The result between 0 copy/reaction and the LoD of ddPCR for each primers/probe sets is defined as suspected SARS-CoV-2 infection, which needs further detection. The outcome 0 copy/reaction for both ORF1ab and N primers/probe sets is judged as negative.

As shown in Figure 3, throat swab samples of each suspected outpatient were firstly collected for laboratory RT-PCR, ddPCR tests and official approved RT-PCR diagnosis in hospitals simultaneously with the same primers/probe sets approved by Chinese CDC (Table 1). Then the suspected outpatients were diagnosed with chest CT. Based on the official medical program of China, outpatients with either official approved RT-PCR (positive) or chest CT (ground glass opacities image, GGO) should be hospitalized. Subsequently, the throat swab samples of all hospitalized patients were collected again and subjected to official approved RT-PCR test at indicated days post hospitalized (Table 1) to monitor the viral load continuously based on the official medical program.

**Fig. 3.**
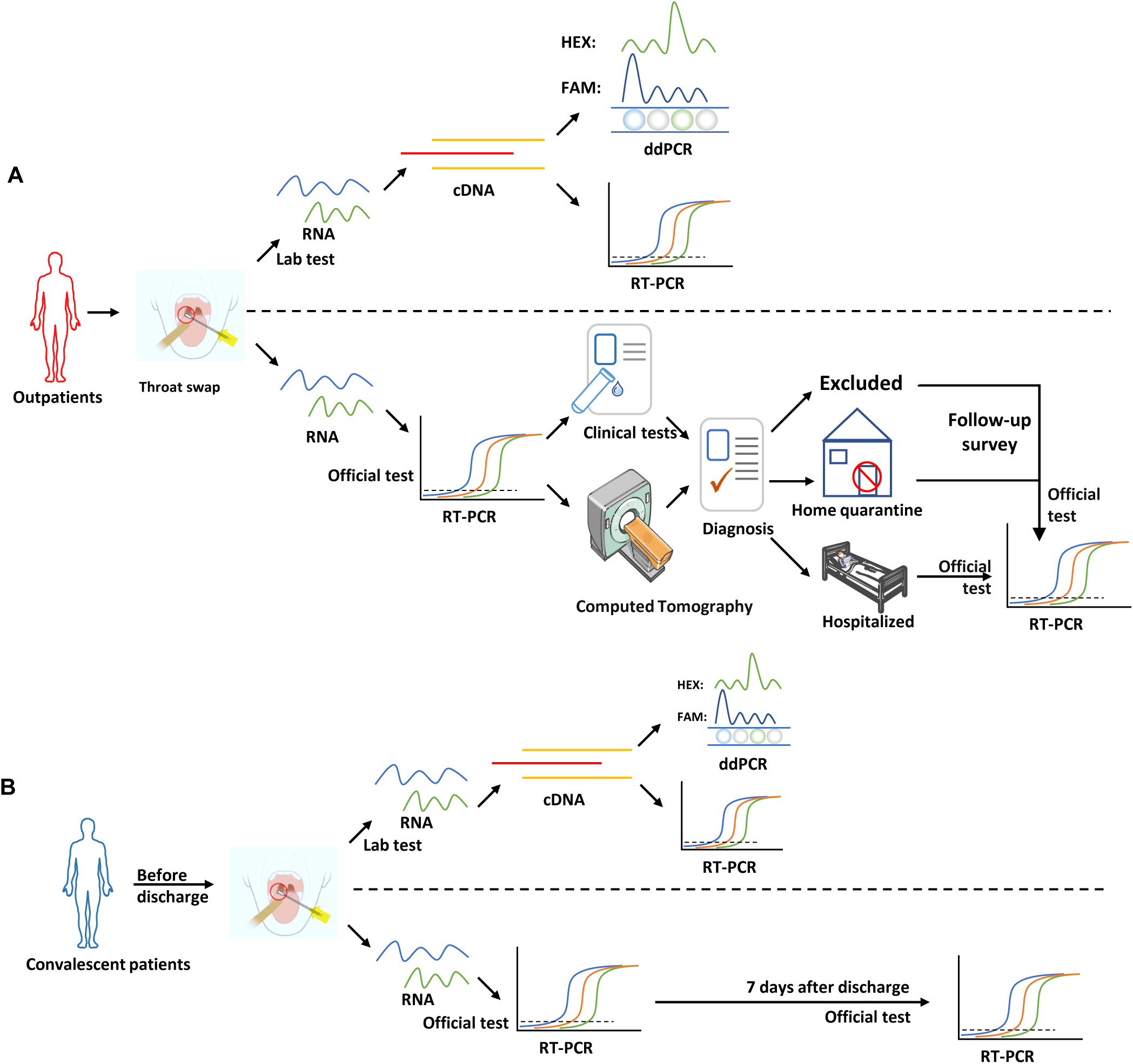
Flowchart of this research design. (A) Research design for suspected outpatients and (B) supposed convalescents. These results were acquired in double-blind from hospitals and laboratory independently. The official approved RT-PCR were conducted by hospitals. The follow-up survey and clinical information of enrolled patients were used to evaluate the performance of ddPCR and RT-PCR.

**Table 1.** RT-PCR and ddPCR Results of febrile and suspected outpatients of COVID-19 and their further clinical information.

| Series No. | Results of RT-PCR in lab (Ct Value)<sup>a, b</sup> |  | Result of ddPCR (copies/reaction)<sup>a, b</sup> |  | Judgment by ddPCR | Initial official reports by RT-PCR<sup>a</sup> | Further diagnosis by chest CT<sup>c</sup> | Patient condition<sup>d</sup> | Official reports by RT-PCR again after (days)<sup>e</sup> |
| --- | --- | --- | --- | --- | --- | --- | --- | --- | --- |
|  | ORF1ab | N | ORF1ab | N |  |  |  |  |  |
| P1 | ND | ND | 0 | 2.0 | P | N | GGO<sup>c</sup> | Hospitalized | P (4d) |
| P2 | 39.46 | 38.90 | 0 | 2.0 | P | N | GGO | Hospitalized | P (4d) |
| P3 | ND | ND | 0 | 2.0 | P | N | GGO | Hospitalized | P (4d) |
| P4 | 35.20 | 35.98 | 4.8 | 20.8 | P | P | GGO | Hospitalized | P (6d) |
| P5 | ND | 39.59 | 0 | 3.6 | P | N | GGO | Hospitalized | P (4d) |
| P6 | 36.24 | 35.45 | 32.4 | 104 | P | P | GGO | Hospitalized | P (6d) |
| P7 | ND | ND | 3.4 | 13.6 | P | N | GGO | Hospitalized | P (2d) |
| P8 | 36.87 | 36.10 | 18.6 | 36.4 | P | P | GGO | Hospitalized | P (6d) |
| P9 | 39.19 | 38.87 | 1.6 | 13.2 | P | N | GGO | Hospitalized | P (3d) |
| P10 | ND | ND | 0 | 1.6 | S | N | GGO | Hospitalized | P (4d) |
| P11 | 35.08 | 33.52 | 4.8 | 16.4 | P | P | GGO | Hospitalized | P (6d) |
| P12 | ND | ND | 0 | 3.6 | P | N | GGO | Hospitalized | P (4d) |
| P13 | ND | ND | 0 | 4.6 | P | N | GGO | Hospitalized | P (4d) |
| P14 | 38.09 | 37.63 | 2.6 | 3.8 | P | N | GGO | Hospitalized | P (2d) |
| P15 | ND | ND | 0 | 3.6 | P | N | GGO | Hospitalized | P (2d) |
| P16 | ND | ND | 0 | 1.8 | P | N | GGO | Hospitalized | P (4d) |
| P17 | 33.97 | 32.76 | 24.4 | 60.8 | P | P | GGO | Hospitalized | P (6d) |
| P18 | ND | ND | 0 | 7.4 | P | N | GGO | Hospitalized | P (2d) |
| P19 | ND | ND | 0 | 1.8 | P | N | GGO | Hospitalized | P (4d) |
| P20 | 37.77 | 36.63 | 16.2 | 36.2 | P | P | GGO | Hospitalized | P (6d) |
| P21 | ND | ND | 3.8 | 1.8 | P | N | GGO | Hospitalized | P (3d) |
| P22 | ND | ND | 2.4 | 0 | P | N | GGO | Hospitalized | P (4d) |
| P23 | 32.19 | 31.81 | 108 | 132 | P | P | GGO | Hospitalized | P (6d) |
| P24 | 28.02 | 27.49 | 1130 | 4440 | P | P | GGO | Hospitalized | P (6d) |
| P25 | ND | ND | 0 | 1.8 | P | N | GGO | Hospitalized | P (4d) |
| P26 | ND | ND | 0 | 1.4 | S | N | GGO | Hospitalized | P (5d) |
| P27 | 25.33 | 24.25 | 544 | 1384 | P | P | GGO | Hospitalized | P (6d) |
| P28 | 27.02 | 26.30 | 22.8 | 98.2 | P | P | GGO | Hospitalized | P (6d) |
| P29 | ND | ND | 0 | 2.2 | P | N | GGO | Hospitalized | P (10d) |
| P30 | 36.15 | 36.02 | 26.2 | 68.4 | P | P | GGO | Hospitalized | P (6d) |
| P31 | ND | ND | 0 | 6.6 | P | N | GGO | Hospitalized | P (8d) |
| P32 | 34.93 | 33.52 | 32.6 | 64.4 | P | P | GGO | Hospitalized | P (6d) |
| P33 | ND | ND | 4.4 | 14.2 | P | N | GGO | Hospitalized | P (7d) |
| P34 | 28.98 | 28.69 | 746 | 1954 | P | P | GGO | Hospitalized | P (6d) |
| P35 | ND | 39.46 | 0 | 3.2 | P | N | GGO | Hospitalized | P (4d) |
| P36 | 31.49 | 30.63 | 36.2 | 140 | P | P | GGO | Hospitalized | P (6d) |
| P37 | 28.22 | 27.26 | 20.8 | 40.4 | P | P | GGO | Hospitalized | P (6d) |
| P38 | 27.09 | 26.48 | 8.6 | 36.2 | P | P | GGO | Hospitalized | P (6d) |
| P39 | ND | ND | 0 | 1.8 | P | N | GGO | Hospitalized | P (8d) |
| P40 | ND | ND | 0 | 3.2 | P | N | GGO | Hospitalized | P (8d) |
| P41 | 32.73 | 33.18 | 16.2 | 11.4 | P | P | GGO | Hospitalized | P (6d) |
| P42 | 34.16 | 33.09 | 22.4 | 32.8 | P | P | GGO | Hospitalized | P (6d) |
| P43 | 25.02 | 27.30 | 1.8 | 2.4 | P | P | GGO | Hospitalized | P (6d) |
| P44 | 37.01 | 36.57 | 16.6 | 28.4 | P | P | GGO | Hospitalized | P (6d) |
| P45 | 35.85 | 35.26 | 10.2 | 32.2 | P | P | GGO | Hospitalized | P (6d) |
| P46 | ND | ND | 0 | 0 | N | N | GGO | Hospitalized | P (6d) |
| P47 | ND | ND | 0 | 0 | N | N | GGO | Hospitalized | P (8d) |
| P48 | ND | 39.27 | 0 | 3.4 | P | N | Pleural bleb | Home Quarantine | NA, lose contact |
| P49 | ND | ND | 0 | 1.2 | S | N | Lower-lobe pneumonia | Home Quarantine | NA, lose contact |
| P50 | ND | ND | 1.6 | 4.0 | P | N | Pneumonia | Home Quarantine | P (10d) |
| P51 | ND | 38.83 | 0 | 3.8 | P | N | Secondary pulmonary tuberculosis | Home Quarantine | P (9d) |
| P52 | 39.83 | 38.51 | 0 | 5.6 | P | N | Normal | Home Quarantine | NA, lose contact |
| P53 | 39.60 | 38.16 | 3.0 | 16.2 | P | N | Fibrous stripes | Home Quarantine | P (7d) |
| P54 | ND | ND | 0 | 2.0 | P | N | Subpleural nodules | Home Quarantine | P (11d) |
| P55 | ND | ND | 0 | 0 | N | N | Normal | Home Quarantine | Negative (4, 6 and 8d), asymptomatic |
| P56 | ND | ND | 0 | 1.2 | S | N | Normal | Excluded | N (6d) |
| P57 | ND | ND | 0 | 0 | N | N | Emphysema | Excluded | NA, lose contact |
| P58 | ND | ND | 0 | 0 | N | N | Fibrous stripes | Excluded | N (6d) |
| P59 | ND | ND | 0 | 0 | N | N | Normal | Excluded | N (5d) |
| P60 | ND | ND | 0 | 0 | N | N | Normal | Excluded | NA, lose contact |
| P61 | ND | ND | 0 | 0 | N | N | Pneumonia | Excluded | N (6d) |
| P62 | ND | ND | 0 | 0 | N | N | Fibrous stripes | Excluded | N (6d) |
| P63 | ND | ND | 0 | 0 | N | N | Nodules | Excluded | N (8d) |

P, positive; N, Negative; S, suspect; ND, not detected; NA, not applicable.
<sup>a</sup> Throat swab samples were firstly collected for laboratory RT-PCR, ddPCR tests and official approved RT-PCR diagnosis in hospitals simultaneously using the same primers/probe sets approved by Chinese CDC. In laboratory tests, the RNA of throat swab samples were extracted and reverse transcribed to cDNA that is subjected to both RT-PCR and ddPCR tests subsequently.
<sup>b</sup> The reaction systems of laboratory RT-PCR and ddPCR are 20 µl/reaction.
<sup>c</sup> The subsequent results of chest CT were used to diagnose the COVID-19 in hospitals based on the official medical program of China. GGO, ground glass opacities image.
<sup>d</sup> Outpatients with chest CT (GGO) or official approved RT-PCR (positive) were hospitalized. Outpatients with chest CT (no GGO), official approved RT-PCR (negative) but ddPCR (positive) were suggested to be quarantined at home. Outpatients with CT (normal), official approved RT-PCR (negative), ddPCR (negative) and other supportive clinical tests (supplementary Table S1) were excluded for COVID-19.
<sup>e</sup> The infection of SARS-CoV-2 was confirmed by official approved RT-PCR test after the indicated days.

The supposed convalescents should be that: (1) temperature returned to normal for more than 3 days, and respiratory symptoms significantly improved; (2) chest CT imaging showed significant absorption of inflammation; (3) the nucleic acid test of respiratory pathogen was negative for two consecutive times, and the sampling interval should be at least 1 day, based on the official medical program. In this study, two throat swab samples of each supposed convalescent (randomly selected) were collected for laboratory RT-PCR, ddPCR and official approved RT-PCR tests simultaneously with the same primers/probe sets (Table 2). Subsequently, the throat swab samples of discharged convalescents were collected again and subjected to official approved RT-PCR test at indicated days post discharge to assess the viral load after discharge (Table 2).

**Table 2.** Results of RT-PCR and ddPCR for supposed convalescents who are about to be discharged after treatments.

| Patient Number | Patient status | Official reports by RT-PCR<sup>a</sup> | Results of RT-PCR in lab (Ct Value)<sup>b, c</sup> |  | Result of ddPCR (copies/reaction)<sup>b, c</sup> |  | Judgment by ddPCR | Official reports by RT-PCR again after (days)<sup>d</sup> |
| --- | --- | --- | --- | --- | --- | --- | --- | --- |
|  |  |  | ORF1ab | N | ORF1ab | N |  |  |
| P64 | Supposed convalescent | N | ND | ND | 0 | 2.4 | P | P (12d) |
| P65 | Supposed convalescent | N | ND | ND | 0 | 2.2 | P | P (7d) |
| P66 | Supposed convalescent | N | 39.33 | 39.05 | 11.4 | 12.0 | P | N (7d) |
| P67 | Supposed convalescent | N | ND | 40.07 | 0 | 9.0 | P | N (6d) |
| P68 | Supposed convalescent | N | 40.03 | 39.82 | 0 | 16.0 | P | N (7d) |
| P69 | Supposed convalescent | N | ND | ND | 1.8 | 0 | S | P (7d) |
| P70 | Supposed convalescent | N | ND | ND | 0 | 2.2 | P | P (7d) |
| P71 | Supposed convalescent | N | 39.79 | 38.90 | 3.8 | 106 | P | P (5d) |
| P72 | Supposed convalescent | N | ND | 40.02 | 1.4 | 1.4 | S | N (7d) |
| P73 | Supposed convalescent | N | ND | ND | 0 | 0 | N | N (7d) |
| P74 | Supposed convalescent | N | 39.61 | ND | 0 | 0 | N | N (7d) |
| P75 | Supposed convalescent | N | ND | 40.01 | 0 | 0 | N | N (7d) |
| P76 | Supposed convalescent | N | ND | ND | 0 | 0 | N | N (7d) |
| P77 | Supposed convalescent | N | ND | ND | 0 | 0 | N | P (7d) |

P, positive; N, Negative; S, suspect; ND, not detected.
<sup>a</sup> Throat swab samples of supposed convalescent were firstly collected for official approved RT-PCR test and laboratory RT-PCR and ddPCR tests.
<sup>b</sup> The collected samples were test for both RT-PCR and ddPCR in laboratory simultaneously using the same reverse transcriptase system and primers/probe sets approved by Chinese CDC.
<sup>c</sup> The reaction systems of laboratory RT-PCR and ddPCR are 20 µl/reaction.
<sup>d</sup> The nucleic acid of SARS-CoV-2 was tested again by official approved RT-PCR after the indicated days to assess the viral load of discharged convalescents.

In the laboratory tests, the RNA of throat swab samples were extracted and reverse transcribed to cDNA that is subjected to both RT-PCR and ddPCR tests. The follow-up survey and clinical information of enrolled cohort were listed in supplementary Table S1 after the laboratory tests.

### Analysis and comparison of the performance of ddPCR and RT-PCR for SARS-CoV-2 diagnosis

Among the 63 suspected outpatients (P1-P63), 21 positive and 42 negative were reported by official approved RT-PCR in two hospitals, which were also double checked by our laboratory RT-PCR (collectively referred to as RT-PCR in performance analysis). In contrast, 49 positive, 10 negative and 4 suspected SARS-CoV-2 infections were reported by ddPCR according to the above criteria (Table 3). The follow-up survey (Table 1 and supplementary Table S1) revealed that 47 cases (P1-P47) out of 63 were hospitalized subsequently with ground glass opacities images (GGO) of chest CT [19], which were further confirmed as SARS-CoV-2 infection by official approved RT-PCR at 2-10 days post hospitalized. Besides, 7 cases (P48-P54) with ddPCR positive (1 suspected), RT-PCR negative and other images (not GGO) of chest CT were suggested to be quarantined at home considering the positive/suspected reports of ddPCR. The follow-up survey revealed that 4 cases (P50, P51, P53 and P54) out of 7 developed difficulty breathing later, and were confirmed as SARS-CoV-2 infection in other hospital. The rest 3 cases (P48, P49 and P52) out of 7 have lost contact for tracking. Of note, 1 case (P55) with ddPCR negative, RT-PCR negative and normal images of chest CT were SARS-CoV-2 IgM/IgG positive. The further tests by official approved RT-PCR still showed negative reports at 4, 6 and 8 days later, indicating asymptomatic infection of SARS-CoV-2. Meanwhile, 6 out of 8 cases (P56-P63) with ddPCR negative (1 suspected), RT-PCR negative and other images (not GGO) of chest CT were excluded by negative reports of SARS-CoV-2 IgM/IgG (P56, P61 and P62) and official approved RT-PCR tests at 5-8 days later (P56, P58, P59 and P61-P63) in the follow-up survey. Moreover, all of the 6 cases have reported good health. The rest 2 cases (P57 and P60) have lost contact for tracking. We further analyzed and compared the performance of RT-PCR and ddPCR for the nucleic acid detection of SARS-CoV-2 according to the follow-up survey and clinical information (Table 4).

**Table 3.**
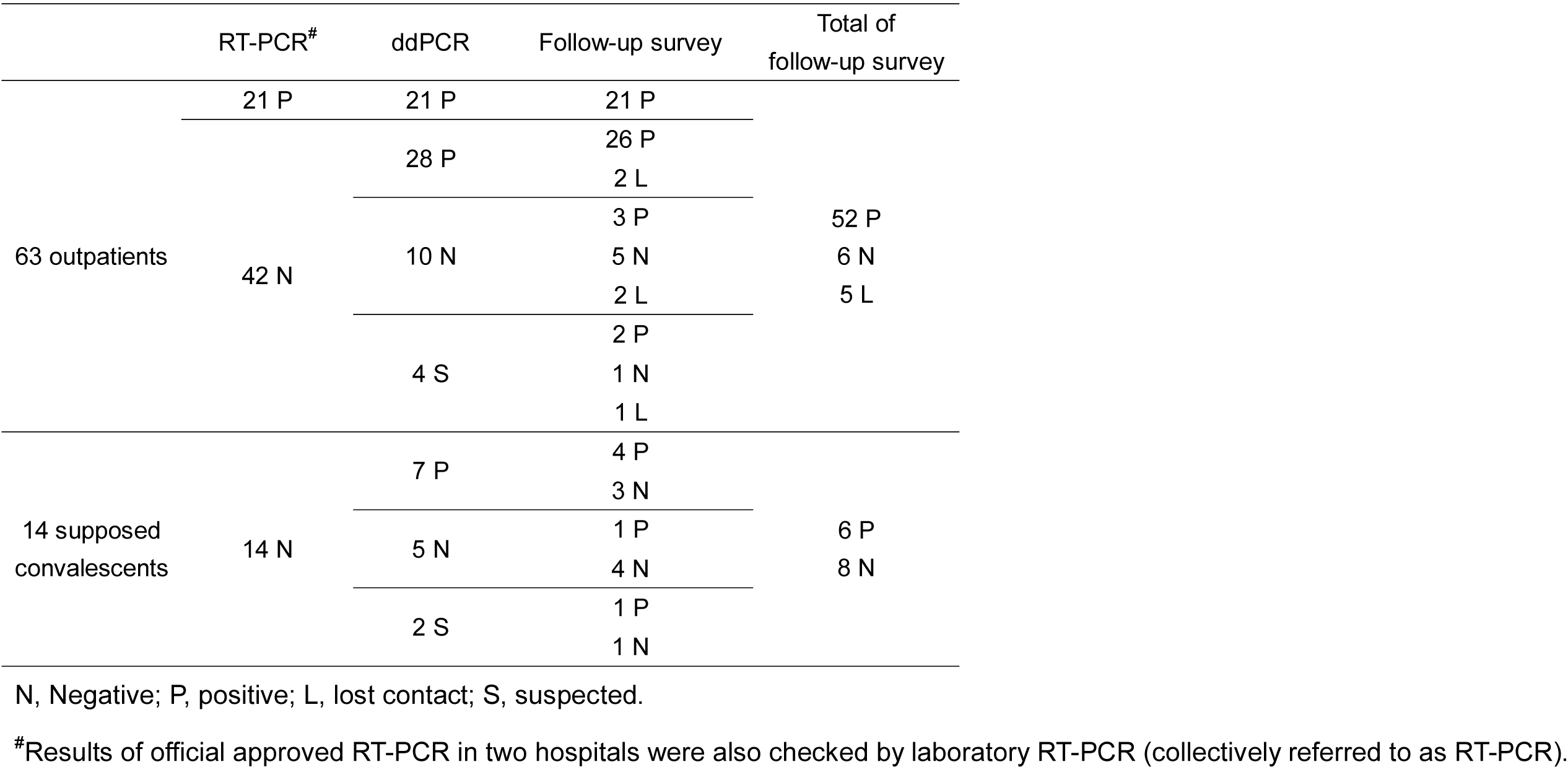
Reports summary of RT-PCR and ddPCR for clinical samples compared to the follow-up survey.

**Table 4.** Diagnostic performance of ddPCR and RT-PCR for suspect patients.

|  |  | Clinically<br>positive | Clinically<br>negative | Sensitivity<br>(95% CI) | Specificity<br>(95% CI) | PPV<br>(95% CI) | NPV<br>(95% CI) | Negative likelihood<br>ratio (95% CI) | Accuracy<br>(95% CI) |
| --- | --- | --- | --- | --- | --- | --- | --- | --- | --- |
| RT-PCR | Test positive | 21 <sup>a</sup> | 0 <sup>b</sup> | 40% | 100% | 100% | 16% | 0.60 | 47% |
|  | Test negative | 31 <sup>c</sup> | 6 <sup>d</sup> | (27-55%) | (54-100%) | (NA) | (13-19%) | (0.48-0.75) | (33-60%) |
| ddPCR | Test positive | 47 <sup>a</sup> | 0 <sup>b</sup> | 94% | 100% | 100% | 63% | 0.06 | 95% |
|  | Test negative | 3 <sup>c</sup> | 5 <sup>d</sup> | (83-99%) | (48-100%) | (NA) | (36-83%) | (0.02-0.18) | (84-99%) |
|  | Suspect | 2 | 1 |  |  |  |  |  |  |

Sensitivity = $[a / (a + c)] \times 100\%$ ; Specificity = $[d / (b + d)] \times 100\%$ ; PPV = $[a / (a + b)] \times 100\%$ , positive predictive value;
NPV = $[d / (c + d)] \times 100\%$ , negative predictive value; Negative likelihood ratio = $(1 - \text{Sensitivity}) / \text{Specificity}$ ;
Accuracy = $[(a + d) / (a + b + c + d)] \times 100\%$ ; NA, not applicable.
Calculated by MEDCALC (https://www.medcalc.org/calc/diagnostic_test.php).

Among the 14 supposed convalescents (P64-P77), whose samples were all reported as negative by official approved RT-PCR, 7 positive, 5 negative and 2 suspected SARS-CoV-2 infections were reported by ddPCR (Table 3). The follow-up survey (Table 2 and supplementary Table S1) revealed that 5 cases (P64, P65 and P69-P71) out of 9 (P64-P72) with ddPCR positive (2 suspected), RT-PCR negative and lesions absorbed images of chest CT have been diagnosed as SARS-CoV-2 nucleic acid positive again by official approved RT-PCR at 5-12 days post discharge. The rest 4 cases (P66-P68 and P72) out of 9 have remained as nucleic acid negative. Meanwhile, another 4 cases (P73-P76) out of 5 (P73-P77) with ddPCR negative, RT-PCR negative and lesions absorbed images of chest CT still remained as nucleic acid negative at 7 days post discharge, indicating functional cure. Of note, 1 case (P77) with the same conditions has returned to nucleic acid positive at 7 days post discharge.

## Discussion

More and more nucleic acid detection kits have been developed for SARS-CoV-2 recently based on RT-PCR to meet the requirement of large-scale clinical molecular diagnosis. It has been reported that 6 kinds of RT-PCR detection kits were compared and analyzed for their detection performance, which showed that there were differences in the detection ability for weakly positive samples, and the accuracy, sensitivity and reproducibility of some kits are not ideal [20]. Meanwhile, other methods, such as chest CT and immunological detection of IgM/IgG have been used to help for the diagnosis of COVID-19. However, the direct detection of virus is irreplaceable. Different from RT-PCR that the data are measured from a single amplification curve and a Cq value, which is highly dependent on reaction efficiency, primer dimers and sample contaminants, ddPCR is measured at reaction end point which virtually eliminates these potential pitfalls.

In this study, we showed that 26 samples from COVID-19 outpatients with RT-PCR negative were detected as positive by ddPCR using the same samples. Accordingly, the NPV of ddPCR (63 %, 36 to 83) is obviously higher than that of RT-PCR (16 %, 13 to 19), which indicates that part of true COVID-19 outpatients (26 positive reports by ddPCR in this study) could not be diagnosed in time by RT-PCR, potentially leading to the higher risk of severe illness and viral spreading (Tables 3 and 4). Remarkably, 4 cases (P50, P51, P53 and P54) with ddPCR positive, RT-PCR negative and other images (not GGO) of chest CT were confirmed as SARS-CoV-2 infection 7–11 days later, indicating that our suggestion of quarantine according to the reports of ddPCR is reasonable. The application of ddPCR for SARS-CoV-2 diagnosis would help to early treatment and control the viral transmission. In conclusion, compared with RT-PCR, ddPCR show superiority for clinical detection of SARS-CoV-2 to reduce the false negatives, which could be a powerful complement to the current standard RT-PCR.

Notably, 6 cases (P64, P65, P69-P71 and P77) out of 14 (42.9 %) supposed convalescent patients, who are negative for throat swab nucleic acid tests twice by RT-PCR, are still carrying SARS-CoV-2 according to the follow-up survey. Although the risk of viral transmission is unknown, the virus is replicating, leading to the increase of viral load. Therefore, the current clinical practice that the convalescent continues to be quarantined for at least 2 weeks is reasonable and necessary. Therefore, we recommend that ddPCR could be a complement to the current standard RT-PCR to re-confirm the convalescent, which would benefit to reduce the risk of the SARS-CoV-2 epidemic and social panic.

However, the specificity and PPV were 100% for both RT-PCR and ddPCR because of no false positives. Partly because the sample size is small, but also the clinical samples we collected were from designated hospitals in Wuhan during the COVID-19 epidemic, which meant the disease prevalence of COVID-19 was higher than common clinical scenarios. Moreover we used only primers/probes sets from China CDC, which could not represent primers from other official institutes. Further research to compare the efficiency of these different primers needs be conducted, which helps to improve the diagnostic accuracy of SARS-CoV-2 detection in different countries.

## Data Availability

The authors confirm that the data supporting the findings of this study are available within the article.

## Author Contributions

YC, KL and JF conceptualized the study design. TS, WH, LD, TC, YX, and GC recruited the patients, collected specimens, collected demographic, clinical data; XL, MG, QZ, XW, YY, MS, DG and ZH did the laboratory tests. JF, YL and QZ plotted the figures; XL, MG, JF and YC analyzed the data; ZH, XK, YL, YLL and YC interpreted the results; JF wrote the initial drafts of the manuscript; YC, JF, HU and KL revised the manuscript and FL and KX commented on it. All authors read and approved the final report.

## Acknowledgements

We are grateful to Beijing Taikang Yicai Foundation for their great support to this work.

## Supplementary information

**Table S1. Clinical information about all patients.**

## References

[1] Wu F, Zhao S, Yu B, et al. A new coronavirus associated with human respiratory disease in China. Nature. 2020;

[2] Chen L, Liu W, Zhang Q, et al. RNA based mNGS approach identifies a novel human coronavirus from two individual pneumonia cases in 2019 Wuhan outbreak. Emerg. Microbes Infect. 2020;9:313–319.

[3] World Health Organization. Laboratory testing for 2019 novel coronavirus (2019-nCoV) in suspected human cases [Internet]. 2020 [cited 2020 Feb 4]. p. 1–7. Available from: https://www.who.int/publications-detail/laboratory-testing-for-2019-novel-coronavirus-in-suspected-human-cases-20200117.

[4] General Office of the National Health and Health Commission O of the SA of TCM. Diagnosis and treatment of pneumonitis with a new type of coronavirus infection (trial version 5) [Internet]. 2020 [cited 2020 Feb 6]. Available from: http://bgs.satcm.gov.cn/zhengcewenjian/2020-02-06/12848.html).

[5] Qiu J. Covert coronavirus infections could be seeding new outbreaks [Internet]. Nat. news. 2020 [cited 2020 Mar 20]. Available from: https://doi.org/10.1038/d41586-020-00822-x.

[6] Wu Z, McGoogan JM. Characteristics of and Important Lessons From the Coronavirus Disease 2019 (COVID-19) Outbreak in China: Summary of a Report of 72 314 Cases From the Chinese Center for Disease Control and Prevention. Jama [Internet]. 2020;2019:25–28. Available from: http://www.ncbi.nlm.nih.gov/pubmed/32091533.

[7] Vogelstein B, Kinzler KW. Digital PCR. Proc. Natl. Acad. Sci. U. S. A. 1999;96:9236–9241.

[8] Pohl G, Shih I-M. Principle and applications of digital PCR. Expert Rev. Mol. Diagn. 2004;4:41–47.

[9] Sanders R, Mason DJ, Foy CA, et al. Evaluation of Digital PCR for Absolute RNA Quantification. PLoS One. 2013;8:e75296.

[10] White RA, Blainey PC, Fan HC, et al. Digital PCR provides sensitive and absolute calibration for high throughput sequencing. BMC Genomics. 2009;10:110–116.

[11] Hindson BJ, Ness KD, Masquelier DA, et al. High-throughput droplet digital PCR system for absolute quantitation of DNA copy number. Anal. Chem. 2011;83:8604–8610.

[12] Hindson CM, Chevillet JR, Briggs HA, et al. Absolute quantification by droplet digital PCR versus analog real-time PCR. Nat. Methods. 2013;10:1003–1005.

[13] Brunetto GS, Massoud R, Leibovitch EC, et al. Digital droplet PCR (ddPCR) for the precise quantification of human T-lymphotropic virus 1 proviral loads in peripheral blood and cerebrospinal fluid of HAM/TSP patients and identification of viral mutations. J. Neurovirol. 2014;20:341–351.

[14] Caviglia GP, Abate ML, Tandoi F, et al. Quantitation of HBV cccDNA in anti-HBc-positive liver donors by droplet digital PCR: A new tool to detect occult infection. J. Hepatol. [Internet]. 2018;69:301–307. Available from: https://doi.org/10.1016/j.jhep.2018.03.021.

[15] Postel M, Roosen A, Laurent-Puig P, et al. Droplet-based digital PCR and next generation sequencing for monitoring circulating tumor DNA: a cancer diagnostic perspective. Expert Rev. Mol. Diagn. 2018;18:7–17.

[16] Miyaoka Y, Mayerl SJ, Chan AH, et al. Detection and Quantification of HDR and NHEJ Induced by Genome Editing at Endogenous Gene Loci Using Droplet Digital PCR. In: Karlin-Neumann G, Bizouarn F, editors. Digit. PCR Methods Protoc. [Internet]. New York, NY: Springer New York; 2018. p. 349–362. Available from: https://doi.org/10.1007/978-1-4939-7778-9_20.

[17] National Institute For viral Disease Control and prevention of PRC. Specific primers and probes for detection 2019 novel coronavirus [Internet]. 2020 [cited 2020 Apr 10]. Available from: http://www.chinaivdc.cn/kyjz/202001/t20200121_211337.html.

[18] Burd EM. Validation of laboratory-developed molecular assays for infectious diseases. Clin. Microbiol. Rev. 2010;23:550–576.

[19] Li M, Lei P, Zeng B, et al. Coronavirus Disease (COVID-19): Spectrum of CT Findings and Temporal Progression of the Disease. Acad. Radiol. [Internet]. 2020;1–6. Available from: https://doi.org/10.1016/j.acra.2020.03.003.

[20] Guo Y., Wang K., Zhang Y., Zhang W., Wang L. LP. Comparison and analysis of the detection performance of six new coronavirus nucleic acid detection reagents. Chongqing Med. 2020;14:1671–8348.

